# Standalone Bio-Interventional Uveoscleral Outflow Enhancement for Intraocular Pressure Reduction in Open-Angle Glaucoma: One-Year Results from a Prospective Multicenter Real-World Evidence Study (NCT05506423)

**DOI:** 10.64898/2025.12.11.25342101

**Authors:** George Reiss, Brian Francis, Quang Nguyen, Reena Garg, Tsontcho Ianchulev, Sandra Sieminski, Paul Singh

## Abstract

This prospective, multicenter, real-world evidence study evaluates the 12-month safety and effectiveness of standalone cyclodialysis with AlloFlo™ cleft reinforcement for intraocular pressure (IOP) reduction in open-angle glaucoma (OAG). AlloFlo represents the world’s first acellular, allogenic scleral tissue implant, and data from this CREST Study cohort (NCT05506423) contribute critical long-term safety and effectiveness knowledge to the field of extracellular matrix biomaterials research, in addition to describing a novel procedure for surgical management of OAG.

Eyes with investigator-confirmed inadequately controlled OAG were treated with standalone cyclodialysis using a microsurgical cannula (CycloPen™), followed by uveoscleral cleft reinforcement with AlloFlo. Eyes were followed prospectively for 12 months. Key outcomes included changes in medicated IOP, number of glaucoma medications, adverse events, and progression to subsequent glaucoma procedures.

Forty-one eyes of 38 patients were included. Most eyes (66%) were considered treatment-refractory, defined as having any of: failed ≥ 1 incisional surgery or cilioablative procedure; condition in which incisional surgery would be more likely to fail than in eyes with uncomplicated OAG. At 12 months, mean IOP decreased 31% to 14.7 ± 6.9 mmHg (within the normal IOP range of 10-20 mmHg, p < 0.001); mean number of glaucoma medications decreased 32% to 1.9 ± 1.6 (p < 0.001). Seventy-one percent of eyes achieved ≥ 20% IOP reduction (a clinically meaningful benchmark set by the FDA). More than half of eyes (53%) achieved ≥ 20% IOP reduction without increasing medication. Three eyes (7.2%) progressed to incisional glaucoma surgery. Postoperative IOP elevations ≥ 10 mmHg occurred in 17% of eyes, most of which resolved within 30 days of the procedure. No persistent inflammation, implant rejection, clinically significant hyphema, or scaffold migration occurred.

These findings suggest that uveoscleral outflow enhancement with AlloFlo provides a safe, conjunctiva-sparing option for IOP reduction in OAG, including eyes with prior surgical interventions.

## Introduction

Glaucoma is the leading cause of irreversible blindness in the United States and worldwide, with primary open-angle glaucoma (POAG) the most common subtype, affecting an estimated 80 million individuals^1^. Despite more than a century of research, intraocular pressure (IOP, pressure exerted from within the eye) remains the only modifiable risk factor demonstrated to slow or stop disease progression. Therefore all glaucoma treatments, whether medical or surgical, aim to reduce IOP — mainly by either reducing aqueous humor production or enhancing aqueous drainage via the uveoscleral or trabecular outflow pathways^2^. Reducing IOP alleviates tension on the optic nerve head (ONH) and slows or stops the progressive neurological degeneration responsible for progressive and permanent vision loss.

Surgical access to the uveoscleral pathway was first introduced in 1905 by Heine with the development of the cyclodialysis procedure, which creates a supraciliary conduit for internal aqueous drainage^3^. Although cyclodialysis was a mainstay surgical technique for many years, it was eventually replaced by trabeculectomy and trans-scleral aqueous drainage implants, which provided more predictable and durable results^4^.

Recent advances in surgical instrumentation and implantable materials, have led to a resurgence of uveoscleral-based surgical approaches^3^. One method for enhancing aqueous outflow involves supraciliary drainage devices for internal stenting and long-term outflow support, though they carry the drawbacks of implantable foreign intraocular hardware. More recently, hardware-free methods, such as the bio-interventional cyclodialysis technique originally described by Ianchulev et al.^5^, have emerged as more bio-compatible alternatives. This approach combines a micro-interventional *ab-interno* cyclodialysis with reinforcement using a micro-trephined allogeneic bio-scaffold. The bio-tissue stabilizes the filtration cleft and supports long-term uveoscleral outflow^6–8^.

This study provides 12-month real-world evidence evaluating the hypothesis that standalone bio-interventional cyclodialysis, followed by cleft reinforcement with acellular allogenic scleral tissue for IOP reduction in eyes with OAG inadequately controlled on medical therapy, including those with prior glaucoma procedures, would lower IOP safety and durably. Safety reporting from this study confirms the compatibility of the world’s first acellular scleral allograft implant, contributing favorable data for future investigations of extracellular matrix (ECM) biomaterials.

## Materials and Methods

### Study Design

This is a 12-month analysis of the consecutive cohort of eyes enrolled in a multicenter, real-world study assessing the safety and effectiveness of the CycloPen and AlloFlo bio-interventional procedure (clinicaltrials.gov identifier NCT05506423). This cohort consists of all eyes that underwent CycloPen + AlloFlo as a standalone procedure with follow-up through 12 months after surgery. Data were collected from participants’ clinical charts and surgery records. The study was conducted under institutional review board (IRB) approval and in accordance with the Declaration of Helsinki. All participants provided written informed consent. The study plan is published on clinicaltrials.gov at https://clinicaltrials.gov/study/NCT05506423#study-plan. Recruitment started on 17 August 2022 and is planned to continue through 31 December 2025.

### Participants

Participants were recruited within each investigator’s clinical practice or via the investigator’s referral network. All eyes in this analysis had investigator-confirmed OAG with open-angle gonioscopy findings and evidence of glaucomatous optic neuropathy, with or without visual field defects. Most eyes had previously undergone ocular surgery for cataract or glaucoma and were inadequately controlled on medical therapy. A CONSORT Diagram is provided in Fig. 1

**Fig 1.**
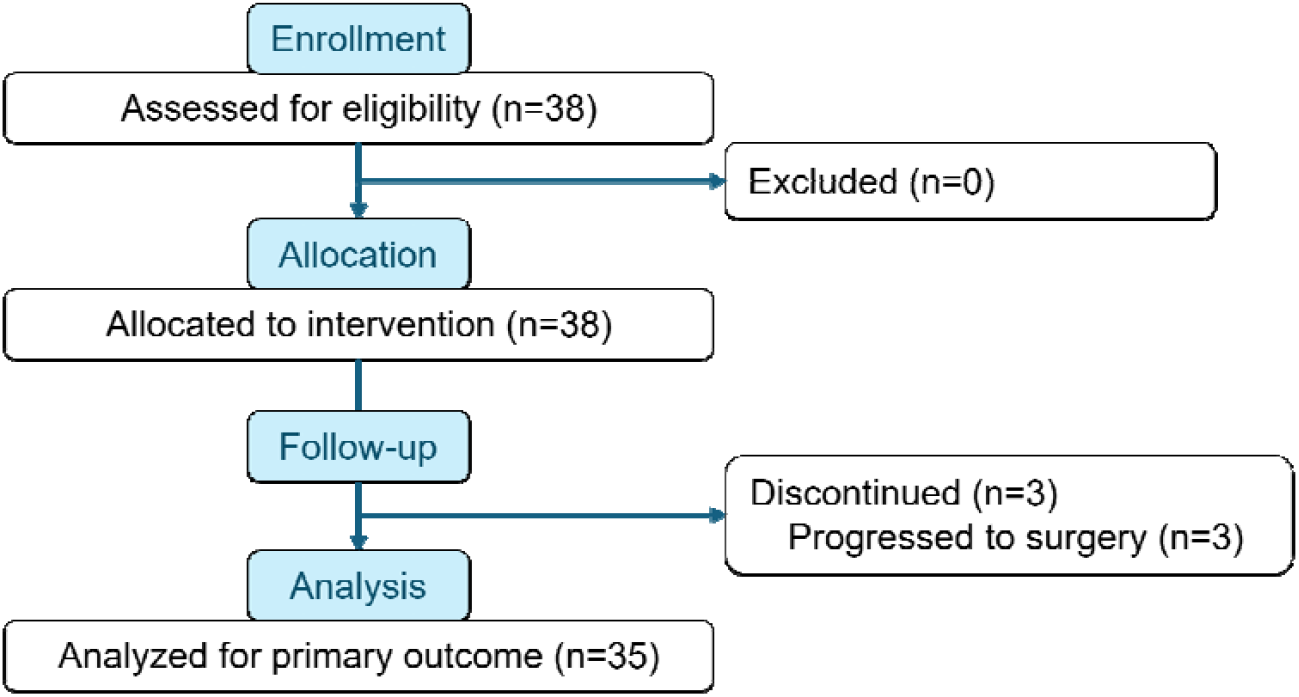
CONSORT 2025 flow diagram.

### Procedure

Following administration of topical anesthesia with optional intracameral lidocaine, sub-tenon, or peribulbar block, gonioscopic visualization of the angle was obtained. A 2 mm temporal corneal incision and subsequent 1.5 mm paracentesis is performed for *ab-interno* access, followed by stabilization of the anterior chamber (AC) with viscoelastic.

The cyclodialysis procedure is performed using a microsurgical cyclodialysis cannula (CycloPen, Iantrek, Inc, White Plains, New York, USA). A 1–2 clock-hour cyclodialysis cleft is created by carefully disinserting the ciliary body from the scleral spur. Viscocycloplasty (augmentation of the cleft with viscoelastic) is then performed to expand the supraciliary filtration reservoir.

The scleral reinforcement procedure is subsequently performed using micro-trephined processed allogeneic bio-tissue (AlloFlo, Iantrek, Inc, White Plains, New York, USA). An elongated 5 mm x 500-micron acellular scleral allograft bio-scaffolding construct is prepared and introduced into the AC under gonio-visualization (Fig 2, Fig 3). The reinforcing spacers are deployed at the endoscleral surface bilaterally and adjacent to the edges of the cyclodialysis. The treating surgeon has discretion to use either one or two allograft spacers, depending on patient physiology. Components do not protrude into the AC, and AC-clear placement is confirmed under direct visualization with the bio-tissue spacers deployed “flush” with the iris root. Any residual viscoelastic is evacuated with irrigation or irrigation/aspiration. Patients were prescribed a surgeon’s choice of postoperative regimen of topical antibiotics and corticosteroids.

**Fig 2.**
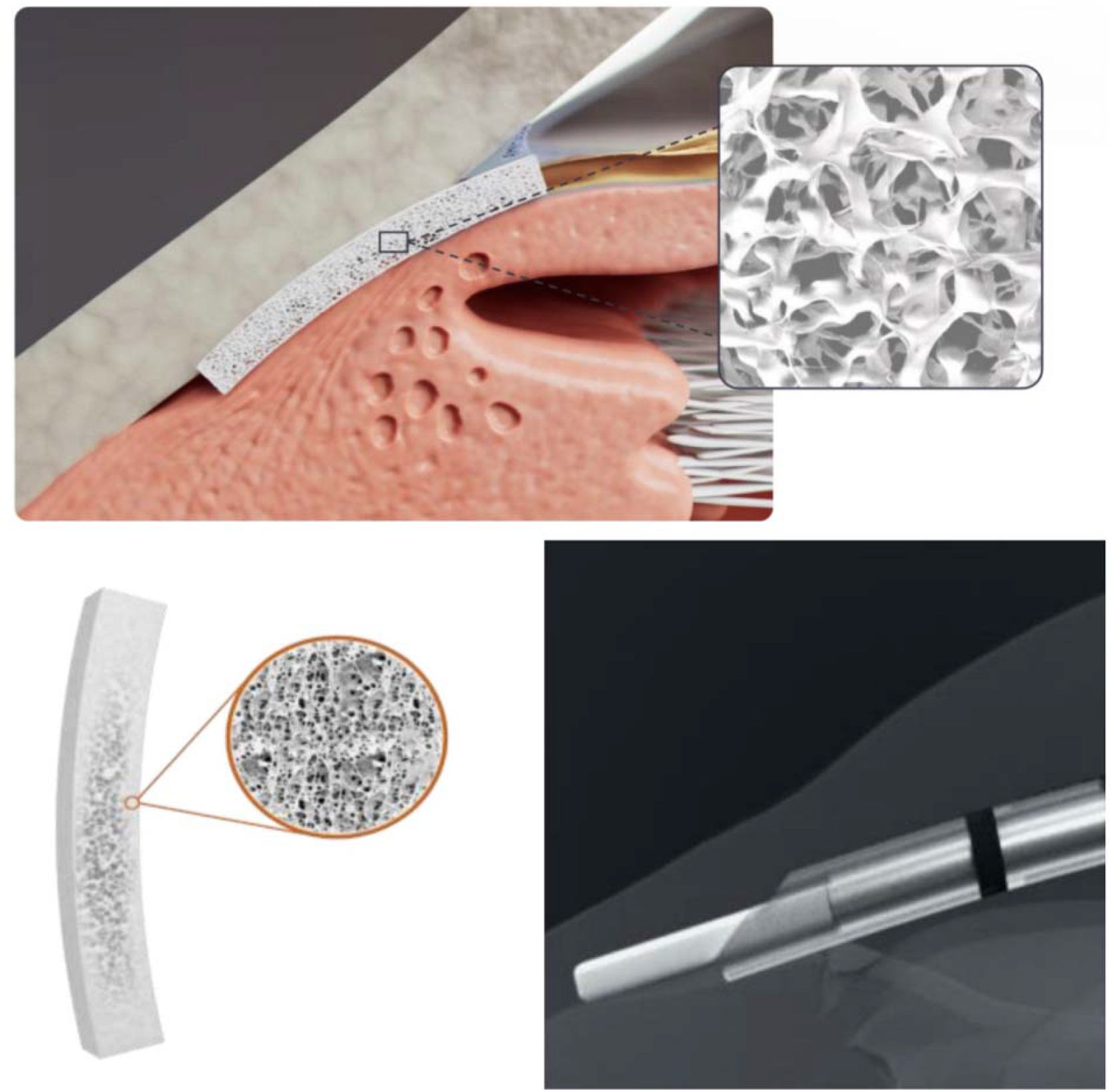
Placement of Allograft Cleft Reinforcement. (Top) Bio-interventional cyclodialysis with the allograft spacer fully deployed to maintain a durable uveoscleral outflow conduit. (Lower-left) processed micro-trephined scleral bio-scaffold made of porous, acellular, hydrophilic, collagenouc sterile matrix. (Lower-right) Sleeved interventional deployment with a cyclodialysis cannula.

**Fig 3.**
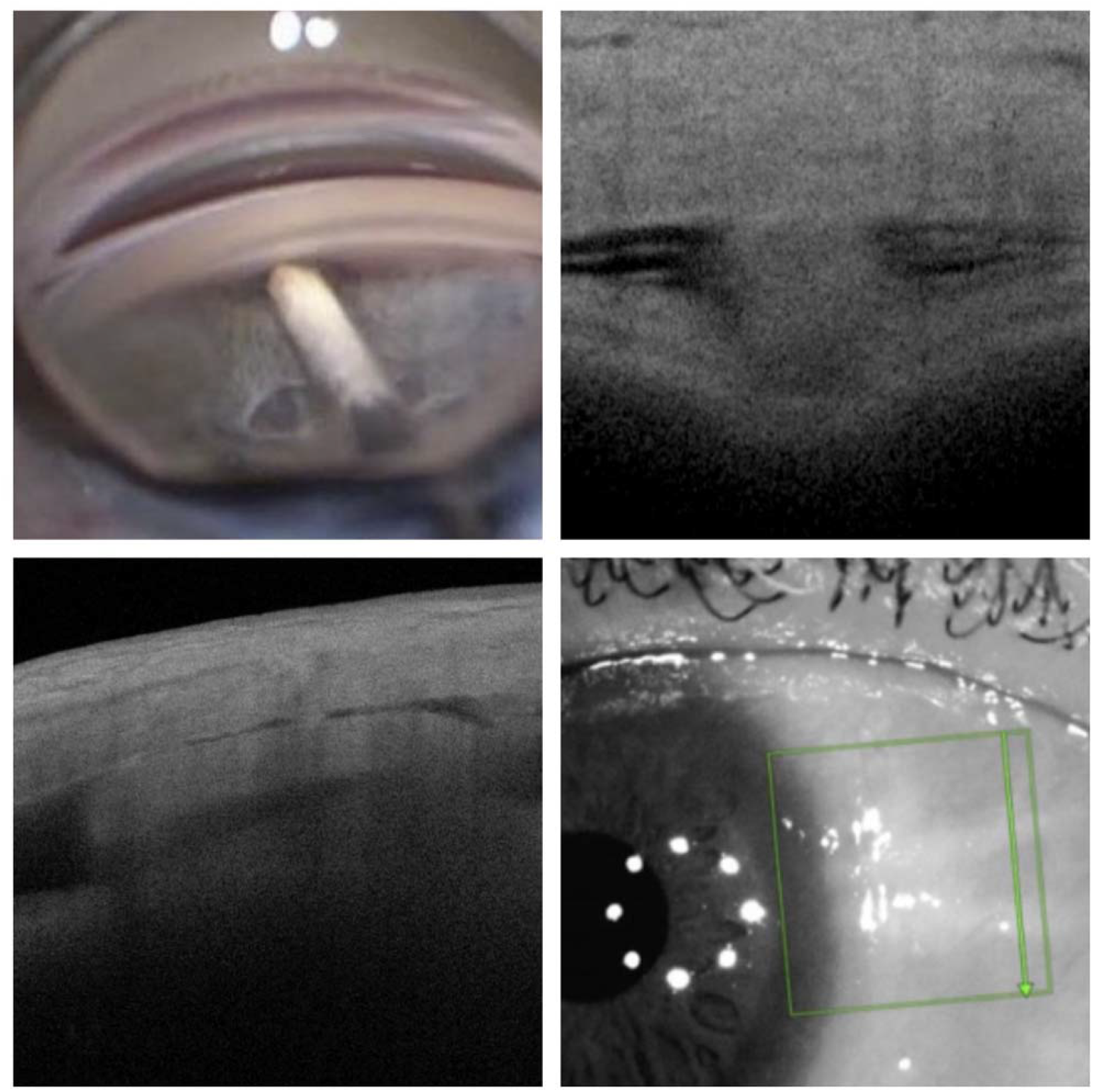
In-situ Images of Allogenic Implant. Clockwise from upper-left: (1) Gonioscopy view of the allogenic bio-spacer during intraocular implantation. (2) Post-implantation 90-day corneal optical coherence tomography (OCT) section of the cyclodialysis with the in-situ allograft reinforcement demonstrating the homologous biologic nature of the tissue in lieu of conventional implantable hardware. (3) Post-implantation 90-day longitudinal OCT imaging of the uveoscleral outflow enhancing bio-scaffold designed to maintain a durable aqueous conduit and filtration reservoir and prevent re-stenosis of the cyclodialysis. (4) External transluminating image demonstrating the visible allograft spacers with the additive reinforcement at 90 days after the procedure.

### Follow-Up and Outcome Measures

Patients were prospectively followed for safety, the need for subsequent glaucoma intervention, corrected distance visual acuity (CDVA), changes in IOP, and changes in glaucoma medications throughout the 12-month postoperative period. Collection of IOP, number of IOP-lowering medications, and CDVA occurred at 1 day, 1 month, 6 months, and 12 months postoperatively.

Key effectiveness outcomes included mean change in medicated IOP and mean change in IOP-lowering medications from baseline to 12 months. Safety assessments included slit lamp and gonioscopy, rate of occurrence of inflammation, clinically significant hyphema, and tissue migration.

### Statistical Analysis

Descriptive statistics were used to summarize continuous (mean ± SD) and categorical data (n, %). Comparisons from baseline to follow-up were made using paired two-tailed t-tests and Fisher’s exact test where appropriate. Significance was defined as two-sided p < 0.05. Qualified success was defined as IOP ≤ 18 mmHg at 12 months without secondary surgery. Results were compiled and programmed in Excel.

## Results

### Patient Demographics and Baseline Disease Characteristics

A total of 41 eyes in 38 patients underwent the standalone uveoscleral outflow procedure with 12-month postoperative follow-up (Table 1). Most eyes (66%) were considered refractory because they had uncontrolled IOP despite maximally tolerated medication therapy (MTMT) or previous incisional surgery or cilioablative procedure. Additionally, most eyes had undergone at least one prior glaucoma procedure: 37% had prior trabecular MIGS, 37% had prior SLT, and 27% had prior incisional glaucoma surgery (e.g., tube shunt, trabeculectomy, gel stent, or cyclophotocoagulation). The mean (± SD) baseline IOP was 21.6 ± 5.0 mmHg with 2.8 ± 1.3 glaucoma medications.

**Table 1.** Patient demographics and baseline disease characteristics.

| **Patient demographics** | **N=38** |
| --- | --- |
| Sex, n (%) female | 17 (44.7%) |
| Age, mean years (± SD) | 71.4 ± 10.5 |
| Race, n (%) |  |
| Asian | 1 (2.6%) |
| Black or African American | 5 (13.2%) |
| White | 32 (84.2%) |
| Hispanic or Latino, n (%) | 13 (34.2%) |
| **Baseline disease characteristics (by eye)** | **N=41<sup>a</sup>** |
| Glaucoma diagnosis, n (%) |  |
| Primary OAG | 38 (92.7%) |
| Secondary OAG | 4 (9.8%) |
| Angle-closure glaucoma | 1 (2.4%) |
| Prior surgical intervention, n (%) |  |
| Cataract surgery/pseudophakic | 28 (68.3%) |
| Canal MIGS (goniotomy, canaloplasty, or stents) | 15 (36.6%) |
| SLT | 15 (36.6%) |
| Incisional glaucoma surgery (e.g., tube shunt, trabeculectomy, CPC) | 11 (26.8%) |
| Baseline mean deviation, mean MD (± SD) | -6.8 ± 5.2 |
| Baseline IOP lowering medications, mean n (± SD) | 2.8 ± 1.3 |
| Refractory (uncontrolled IOP with MTMT or incisional surgery), n (%) | 27 (65.9%) |

IOP, intraocular pressure; OAG, open-angle glaucoma; MD, mean deviation; MIGS, minimally invasive glaucoma surgery; MTMT, maximally tolerated medication therapy; SD, standard deviation; SLT, selective laser trabeculoplasty.
a. Three patients had bilateral placement of AlloFlo allogenic scleral scaffold; eyes could have multiple diagnoses.

### IOP and Medication Outcomes

At 12 months after the study procedure, eyes had a 31% reduction in mean IOP, reduced from a mean (± SD) of 21.6 ± 5.0 mmHg at baseline to 14.7 ± 6.9 mmHg (p < 0.001; Table 2). Eyes also had a 32% reduction in glaucoma medications, reduced from a mean (± SD) of 2.8 ± 1.3 agents at baseline to 1.9 ± 1.6 (p < 0.001). Importantly, 71% of eyes had a clinically relevant (≥ 20%) IOP reduction from baseline (a clinically meaningful benchmark established by the FDA as an endpoint in glaucoma device clinical trials^9^), and approximately half of eyes (53%) achieved this target without increasing medication.

**Table 2.** Effectiveness outcomes.

| **IOP outcomes** | **N=41** |
| --- | --- |
| Baseline IOP, mean mmHg (± SD), N=39 | 21.6 ± 5.0 |
| 1-month IOP, mean mmHg (± SD), N=39 | 15.6 ± 8.4 |
| 6-month IOP, mean mmHg (± SD), N=32 | 16.8 ± 8.5 |
| 12-month IOP, mean mmHg (± SD), N=31 | 14.7 ± 6.9 |
| Change from Baseline at Month 12, mean mmHg (± SD) | -6.7 ± 6.1 |
| % change from Baseline at Month 12, mean | -31% |
| p-value for change | < 0.01 |
| ≥ 20% IOP reduction from Baseline, % | 71% |
| ≥ 20% IOP reduction from Baseline without increasing medication, % | 53% |
| **Medication outcomes** | **N=41** |
| Baseline, mean number of glaucoma medications (± SD), N=39 | 2.8 ± 1.3 |
| 1 month, mean number of glaucoma medications (± SD), N=39 | 1.7 ± 1.6 |
| 6 months, mean number of glaucoma medications (± SD), N=32 | 1.9 ± 1.5 |
| 12 months, mean number of glaucoma medications (± SD), N=31 | 1.9 ± 1.6 |
| Change from Baseline, mean number of glaucoma medications (± SD) | -1.0 ± 1.2 |
| % change from Baseline at Month 12, mean | -32% |
| p-value for change | < 0.01 |
| No. of glaucoma medications at 12M, n (%) |  |
| 0 | 7 (22.6%) |
| 1 | 9 (29.0%) |
| 2 | 8 (25.8%) |
| 3+ | 11 (35.5%) |
| **12-Month responder analysis, n %** | **N=34<sup>a</sup>** |
| Responder through 12 Months<sup>b</sup> | 28 (82%) |
| 12-month IOP ≤ 21 mmHg | 27 (79%) |
| 12-month IOP ≤ 18 mmHg | 21 (62%) |
| 12-month IOP ≤ 15 mmHg | 18 (53%) |
| Responder & ≥ 20% IOP reduction & no medication increase | 18 (53%) |
| Responder & ≥ 20% IOP reduction & no medication increase & Baseline IOP ≥ 16 (N=29) | 19/29 (66%) |

IOP, intraocular pressure; MIGS, minimally invasive glaucoma surgery; SD, standard deviation.
a. N=34 eyes with data through 12 months of follow-up; three patients who progressed to incisional surgery counted as failures on the responder analysis and not reported in IOP and medication outcomes.
b. Responder defined as eye that had not progressed to incisional glaucoma surgery (i.e., non-MIGS procedure) through 12 months of follow-up.

IOP reductions were consistent among eyes that had undergone trabecular MIGS or SLT prior to uveoscleral outflow enhancement and cleft reinforcement (Table 3). In eyes with prior MIGS (N=10), mean (± SD) IOP decreased 6.5 mmHg (from 21.3 ± 2.9 mmHg at baseline to 14.4 ± 0.8 mmHg). In eyes with prior SLT (N=11), mean (± SD) IOP decreased 6.1 mmHg (from 21.3 ± 5.2 mmHg at baseline to 14.8 ± 4.9 mmHg.

**Table 3.** Effectiveness outcomes by prior procedure.

| **Eyes with prior trabecular MIGS\*** | **N=11** |
| --- | --- |
| IOP outcomes, mean mmHg (± SD) | |
| Baseline IOP | 21.3 ± 5.9 |
| 12-month IOP | 14.4 ± 6.2 |
| Change from Baseline at Month 12 | -6.5 ± 3.8 |
| % change from Baseline at Month 12, mean | -30% |
| Medication outcomes, mean number of glaucoma medications (± SD) | |
| Baseline | 2.7 ± 2.6 |
| 12-month medications | 2.9 ± 2.1 |
| Change from Baseline at Month 12 | -0.3 ± 1.2 |
| % Difference between the means | -11% |
| **Eyes with prior SLT\*** | **N=10** |
| IOP outcomes, mean mmHg (± SD) | |
| Baseline IOP | 21.3 ± 5.8 |
| 12-month IOP | 14.8 ± 6.5 |
| Change from Baseline at Month 12 | -6.1 ± 3.8 |
| % change from Baseline at Month 12, mean | -28% |
| Medication outcomes, mean number of glaucoma medications (± SD) | |
| Baseline | 2.7 ± 1.6 |
| 12-month medications | 2.8 ± 1.3 |
| Change from Baseline at Month 12 | -0.2 ± 1.2 |
| % Difference between the means | -7% |

IOP, intraocular pressure; MIGS, minimally invasive glaucoma surgery; SD, standard deviation; SLT, selective laser trabeculoplasty.
\* Three eyes had prior trabecular MIGS and SLT at baseline.

### Safety Outcomes

The procedure demonstrated a favorable safety profile, with no reported cases of device migration, no persistent or severe postoperative inflammation, no adverse events related to the allogenic bio-scaffold, no implant rejection, and no hyphema or vision-threatening complications (Table 4). There was a single case of same-day anteriorization of the implant, which was also seen on postoperative day 1 and subsequently corrected with proper deployment and good postoperative stability.

**Table 4.**
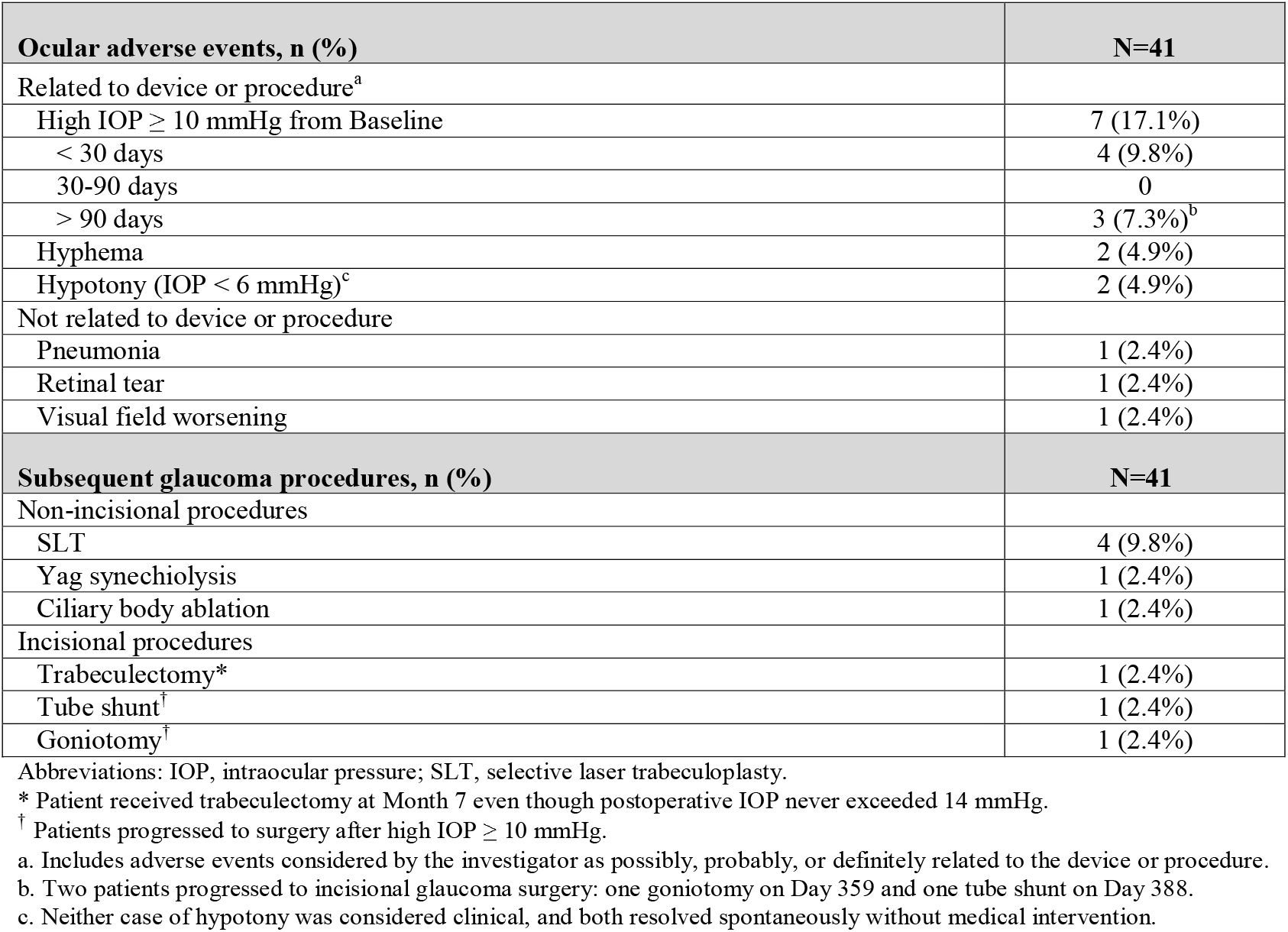
Post-surgical safety.

| Ocular adverse events, n (%) | N=41 |
| --- | --- |
| Related to device or procedure<sup>a</sup> |  |
| High IOP ≥ 10 mmHg from Baseline | 7 (17.1%) |
| < 30 days | 4 (9.8%) |
| 30-90 days | 0 |
| > 90 days | 3 (7.3%)<sup>b</sup> |
| Hyphema | 2 (4.9%) |
| Hypotony (IOP < 6 mmHg)<sup>c</sup> | 2 (4.9%) |
| Not related to device or procedure |  |
| Pneumonia | 1 (2.4%) |
| Retinal tear | 1 (2.4%) |
| Visual field worsening | 1 (2.4%) |
| **Subsequent glaucoma procedures, n (%)** | **N=41** |
| Non-incisional procedures |  |
| SLT | 4 (9.8%) |
| Yag synechiolysis | 1 (2.4%) |
| Ciliary body ablation | 1 (2.4%) |
| Incisional procedures |  |
| Trabeculectomy* | 1 (2.4%) |
| Tube shunt<sup>†</sup> | 1 (2.4%) |
| Goniotomy<sup>†</sup> | 1 (2.4%) |

Abbreviations: IOP, intraocular pressure; SLT, selective laser trabeculoplasty.
\* Patient received trabeculectomy at Month 7 even though postoperative IOP never exceeded 14 mmHg.
<sup>†</sup> Patients progressed to surgery after high IOP ≥ 10 mmHg.
a. Includes adverse events considered by the investigator as possibly, probably, or definitely related to the device or procedure.
b. Two patients progressed to incisional glaucoma surgery: one goniotomy on Day 359 and one tube shunt on Day 388.
c. Neither case of hypotony was considered clinical, and both resolved spontaneously without medical intervention.

The need for subsequent incisional glaucoma surgery (Figure 4) was low: only 3 eyes (7.2%) required incisional surgery through the 12-month follow-up, all due to inadequate IOP control, which included one patient whose IOP increased ≥ 10 mmHg from baseline. No subsequent surgeries were due to complications of the index procedure. There were no reports of clinical hypotony. Postoperative IOP elevation ≥ 10 mmHg from baseline occurred in 7 eyes (17%), most of which (4 of 7) occurred within 30 days of the procedure and resolved spontaneously with conservative medical therapy, no requiring incisional surgery. One of these eyes received subsequent SLT on Day 44. The other three cases occurred > 90 days after surgery — one did not lead to subsequent surgery, one led one led to ciliary body ablation at Month 3, and one led to implantation of a tube shunt at Month 12.

**Fig 4.**
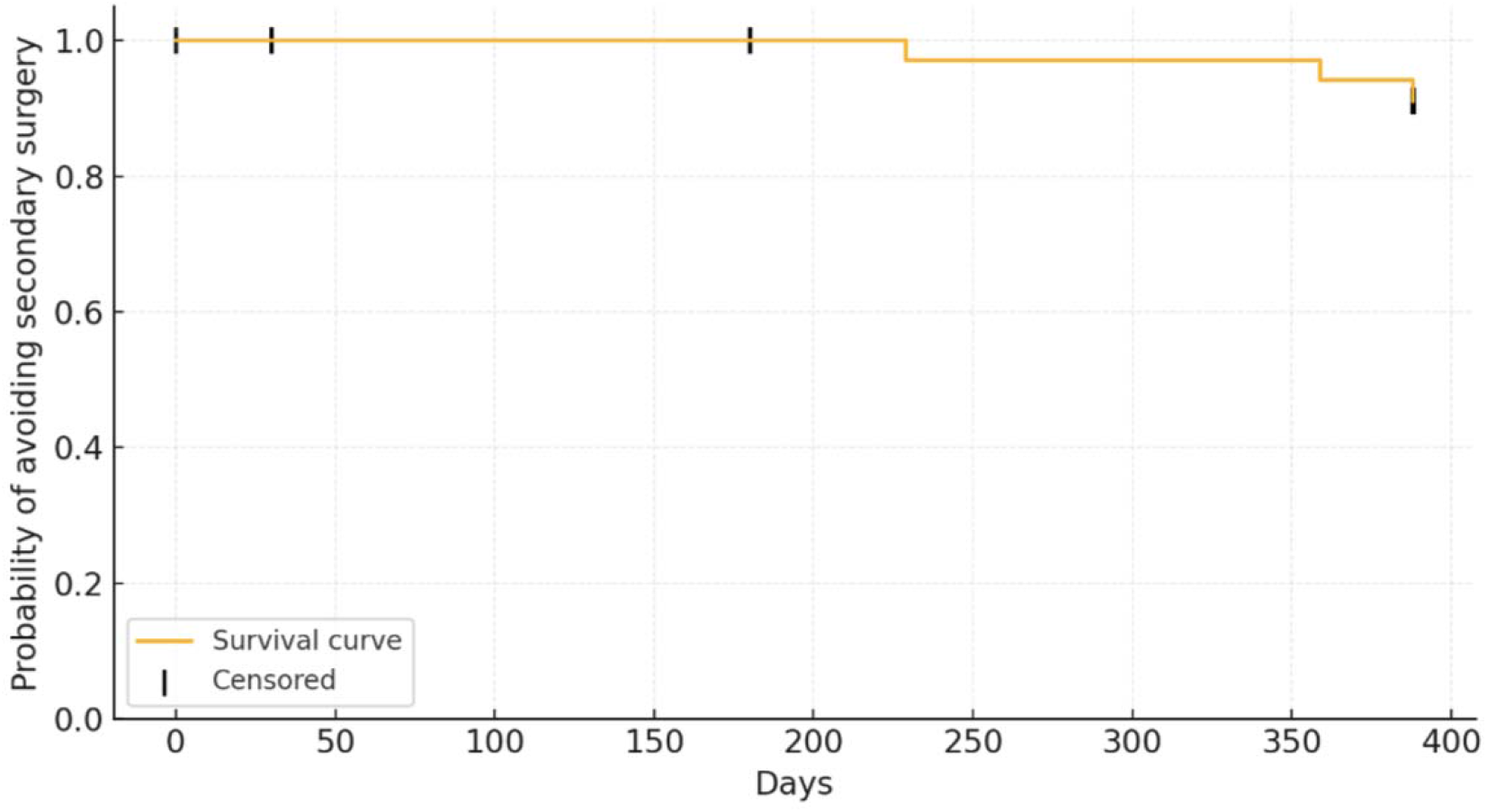
Kaplan-Meier estimate of surgery-free survival after index procedure. Note: N=41 eyes. The curve displays the cumulative probability that an eye remained free of subsequent incisional glaucoma surgery (e.g., tube shunt, trabeculectomy, gel stent, or cyclophotocoagulation) among the 41 treated eyes. 100% of eyes remained free of subsequent surgery through 6 months, 97% at 229 days, 94% at 359 days, and 91% at the end of follow-up (388 days).

## Discussion

This study results demonstrate statistically significant and clinically meaningful IOP lowering after standalone bio-interventional uveoscleral outflow enhancement in patients who are inadequately controlled with topical medication therapy, previous glaucoma procedures, or both. Reduction in mean IOP and number of IOP lowering medications was immediate and sustained over the entire 12-month follow-up period across primary and secondary efficacy outcomes. As a standalone treatment, allograft-reinforced cyclodialysis achieves a clinically meaningful and durable augmentation of uveoscleral outflow with a favorable interventional risk-benefit profile, significantly reducing the need for more invasive, bleb-forming surgeries (e.g., trabeculectomy and tubes).

The magnitude of IOP reduction was notable: 31% mean IOP reduction with 82% of eyes achieving responder status through 12 months. Moreover, the need for escalation to filtering or tube-based procedures was significantly reduced. While 66% of eyes were surgical candidates at baseline due to uncontrolled IOP on maximal medical therapy, only 7.2% required further incisional intervention through the 12-month follow-up. In addition, nearly 80% of eyes achieved IOP control (defined as < 21 mmHg) without increasing medications.

These findings are consistent with prior reports of bio-interventional cyclodialysis performed in combination with cataract surgery. However, the present study isolates the IOP-lowering effects of the procedure alone, eliminating the confounding hypotensive effect of phacoemulsification after cataract surgery, as described in previous reports. The results also parallel those of other uveoscleral-targeting approaches, such as suprachoroidal stents, which have demonstrated high drainage potential and long-term efficacy.

These results of bio-interventional uveoscleral outflow enhancement compare favorably with published results of stented trabecular outflow enhancement with iStent and Hydrus in stand-alone cases^10^. In the COMPARE standalone trial, there was only 1.1 and 1.7 mmHg IOP reduction from medicated baseline for iStent and Hydrus at 12 months, respectively, compared to 6.7 mmHg with the uveoscleral outflow procedure. Change in mean IOP lowering medication burden at 12 months was 1.0 and 1.6 for iStent and Hydrus, respectively, compared to 1.0 for the uveoscleral procedure — from similar pre-operative baselines of 2.5-2.7 medications.

The rate of secondary incisional surgery in the COMPARE trial was 2.6% in the iStent group while in the more advanced CREST population the rate of secondary incisional surgery was 7.2%. Patients enrolled in the current study cohort had more advanced glaucoma with a baseline medicated IOP of 21.6 mmHg vs 19.1 mmHg in the COMPARE trial.

Furthermore, uveoscleral outflow enhancement results compare favorably with the iStent infinite trial where three iStents are implanted in patients with advanced OAG^11^. There was a mean IOP reduction of 5.5 mmHg at 12 months from a medicated baseline of 23.5 mmHg compared to the reduction in this cohort of 6.7 mmHg from a medicated baseline of 21.6 mmHg. The rate of secondary surgical intervention for IOP control in the iStent study was 4.2%.

Overall, the safety of newer interventional procedures for uveoscleral outflow enhancement seems to be closing the gap by demonstrating canal MIGS-like safety profile, while tapping the higher therapeutic index of the uveoscleral drainage pathway. This provides a natural next-generation technological advancement of the glaucoma interventional paradigm to raise the efficacy bar similar to the pharmacologic treatment parallel where uveoscleral outflow drugs represent the mainstay category with the highest efficacy. A recent meta-analysis confirms the surgical parallel of superior uveoscleral outflow biologic effect over trabecular outflow enhancement: at 24 months, there was a greater reduction in IOP from baseline in the uveoscleral interventional group vs trabecular stenting (two iStents; −9.57 vs. −4.92 mmHg, p=0.03). The change from baseline in mean medication use was −1.00 medications with MINIject and −0.56 medications with iStent^12^.

The clinical utility of uveoscleral intervention can be significant, as millions of glaucoma patients have already exhausted the canal interventional option with stents or goniotomy and canaloplasty and are facing waning efficacy with the prospects of highly invasive bleb-forming surgery. Uveoscleral intervention can open the second native/endogenous outflow pathway of higher outflow capacity to delay or diminish the need for more invasive bleb-based procedures, such as trabeculectomy or tubes. Currently, less than 10% of trabecular outflow canal-based MIGS procedures, such as iStent and Hydrus, are used for standalone glaucoma intervention outside being an adjunct to cataract surgery due to the limited efficacy of the trabecular outlow pathway. Even with three iStent implants, the Infinite pivotal trial reported only 23% IOP reduction (5.5 mmHg) using^13^, whereas the single uveoscleral intervention in the present study demonstrated 32% reduction (6.7 mmHg).

Safety outcomes were favorable and consistent with those reported for other minimally invasive interventions. Only three eyes (7.2%) required subsequent incisional glaucoma intervention, and IOP spikes ≥ 10 mmHg occurred in only 17% of cases, predominantly during the postoperative steroid taper period. Given the known steroid sensitivity in up to one-third of glaucomatous eyes^14^, these elevations were expected and largely manageable; nearly half resolved spontaneously without surgical or pharmacologic escalation. This incidence is comparable to rates reported for other procedures, including 17% with gonioscopy-assisted transluminal trabeculotomy (GATT), 21.5% with Xen Gel Stent, and 32% with trabeculotomy^15–17^. The postoperative safety results are not dissimilar to the standalone results reported with iStent Infinite, in which 22% of eyes had significant postoperative IOP increase and 11.1% underwent secondary glaucoma surgery^13^.

No cases of scaffold migration, extrusion, or persistent tissue-related complications were observed. Mild hyphema occurred in only 4.9% of eyes, resolving spontaneously without clinical intervention. The favorable safety profile is likely attributable to the implant’s biologic, acellular composition and the intra-cleft deployment that avoids contact with the corneal endothelium. The collagenous matrix is engineered to mimic native scleral biomechanics while promoting aqueous permeability and minimizing inflammation, fibrosis, or immune reaction.

This study is one of the few to provide clinical outcomes from standalone interventional glaucoma treatment. Nevertheless, the study is limited in scope and duration with only 41 eyes and 12-month outcomes. The relatively small sample size limits subgroup analyses and generalizability. As a real-world evidence investigation, it reports true real-world effectiveness and safety, rather than effectiveness in a rigid and selective protocol design. Because of this, there is inherent variability in treatment and patient selection which captures the standard of care clinical paradigm. There was no prescribed medication re-introduction procedure and there was no medication wash-out. Post-surgical medication regimen was also non-standardized and physician specific.

While the study is real-world evidence, unlike passive retrospective registries, the postoperative follow-up was prospective, monitored, and consistent with Good Clinical Practice (GCP) where patients were consented, and investigators followed the investigational protocol for safety and efficacy outcomes. There was minimal attrition in patient follow-up, and the study was inclusive of a high-risk, real-world glaucoma population, including patients with prior failed surgical interventions.

## Conclusion

Standalone bio-interventional cyclodialysis reinforced with an allogeneic scleral scaffold offers a safe and effective method for sustained IOP reduction in patients with open-angle glaucoma. The procedure may delay or reduce the need for more invasive filtering or tube surgeries, particularly in cases where prior trabecular MIGS have failed.

## Data Availability

All data produced in the present work are contained in the manuscript

## Author Contributions

- *George Reiss MD - writing the report, analyzing data, interpreting results*
- *Brian Francis MD, Doheny Eye Institute, UCLA, Los Angeles, CA-writing the report, analyzing data, interpreting results*
- *Quang Nguyen MD, Scripps Clinic, San Diego, CA - writing the report, analyzing data, interpreting results*
- *Reena Garg MD Georgetown University Hospital, Washington DC - writing the report, analyzing data, interpreting results*
- *Tsontcho Ianchulev MD MPH, New York Eye and Ear of Mount Sinai, New York – writing the report, analyzing data, interpreting results*
- *Sandra Sieminski MD, Ross Eye Institute, Buffalo, NY - writing the report, analyzing data, interpreting results*
- *Paul Singh MD, The Eye center of Racine and Kenosha, Wisconsin - writing the report, analyzing data, interpreting results*

